# Leveraging AI and Transfer Learning to Enhance Outcome Prediction for Out-of-Hospital Cardiac Arrest in Diverse Settings: Insights from the Pan-Asian Resuscitation Outcomes Study

**DOI:** 10.1101/2025.05.11.25327388

**Authors:** Siqi Li, Yohei Okada, Wenjun Gu, Michael Hao Chen, Son Ngoc Do, Pham Dinh Quyet, Quoc TA Hoang, Marcus Eng Hock Ong, Nan Liu, the PAROS Investigators

## Abstract

**Background:** Access to trustworthy artificial intelligence (AI) models for clinical applications like emergency care is unevenly distributed globally due to healthcare inequities. Low-resource settings face challenges in AI model development due to limited data, small sample sizes, and inconsistent data quality. Moreover, models developed from high-resource settings are often not readily applicable in low-resource contexts. Transfer learning (TL) is an AI technology that adapts established models to new settings and offers a potential solution. This study explores the feasibility of TL in clinical contexts, using neurological outcome prediction for out-of-hospital cardiac arrest (OHCA) as a proof of concept.

**Methods:** The Pan-Asian Resuscitation Outcomes Study (PAROS) network provides a multicenter registry for OHCA across the Asia-Pacific region. We applied TL to adapt a neurological outcome prediction model for OHCA, originally developed using a large Japanese cohort (i.e., the external model), to two PAROS registry countries: Vietnam (243 patients) and Singapore (15,916 patients). Separate TL models, calibrated with local data from Vietnam or Singapore, were developed and compared with the external model. Their predictive performance was then compared with that of the external model.

**Findings:** The external model performed poorly on the Vietnam cohort, with an area under the receiver operating characteristic curve (AUROC) of 0·467 (95% CI: 0·141-0·785). The TL-Vietnam model significantly improved performance (AUROC = 0·807, 95% CI: 0·626-0·948). In Singapore, the TL-Singapore model demonstrated modest improvements (AUROC = 0·955, 95% CI: 0·940–0·967), up from 0·945 (95% CI: 0·929–0·958) in the external model.

**Interpretation:** This study highlights the potential of TL to improve prediction accuracy in low-resource settings worldwide, promoting global healthcare equity.

**Funding:** This study was supported by SingHealth Duke-NUS ACP Programme Funding, National Medical Research Council, Clinician Scientist Awards, Ministry of Health, Health Services Research Grant, Singapore, and Laerdal Foundation.

## Introduction

Artificial Intelligence (AI) has become a powerful tool in supporting healthcare and clinical decision-making^1^, but its benefits are unevenly distributed due to significant healthcare inequities^2^. Much of health AI research has focused on high-income and upper-middle-income countries, raising concerns about the generalizability and applicability of AI solutions globally, particularly in low-resource settings^2^. Zyl et al.^3^ identified financial constraints as one of several factors contributing to challenges in low-data-resource settings (LDRS)^3,4^, where disparities are further compounded by systemic issues beyond financial pressures. This results in more pronounced healthcare inequities than reported in studies focusing solely on financial barriers, as the assumption that healthcare solutions in high-income countries can address all population needs may be unfounded^3^.

The demand for healthcare services worldwide is increasing due to factors like population aging and the growing complexity of medical conditions. In response, healthcare systems worldwide must find innovative strategies to manage limited resources effectively. Risk assessment, which prioritizes patients at higher risk of poor outcomes or requiring urgent intervention, is central to this process. However, the development of accurate and context-specific risk prediction models in LDRS is constrained by limited access to large, high-quality datasets, preventing the creation of tailored solutions for these populations.

In such settings, clinicians frequently rely on externally developed models trained on high-resource datasets^5^. However, these models often experience significant performance declines when applied to populations that differ from the original dataset^6^. For example, the HAS-BLED score^7^, a tool to assess bleeding risk in atrial fibrillation patients, was developed using a European cohort but dropped significantly in performance when validated in more diverse global cohorts^8^. Specifically, the area under the receiver operating characteristic curve (AUROC) decreased from 0·72 (95% CI: 0·65-0·79) to 0·65 (95% CI: 0·61–0·68)^6–8^, highlighting the need for adaptable AI models that can be localized to specific regions and populations.

Transfer learning (TL) is an advanced AI technique designed to adapt pre-trained models, built on large datasets, to new settings with limited local data^9^. As illustrated in Figure 1, this technique enables the reuse of existing model parameters to improve performance in new settings without the need for extensive new data collection^10^, making it particularly valuable in LDRS. For example, Hwang et al.^11^ demonstrated the effectiveness of TL by adapting an existing deep neural network model^12^, developed from the Korean National Health and Nutritional Examination Survey data, to predict low-density lipoprotein cholesterol levels, further refining it with local data from Wonju Severance Christian Hospital^11^. Despite its potential, the application of TL in clinical research, particularly in LDRS, remains underexplored, presenting an opportunity to address critical gaps in global healthcare equity^10^.

**Figure 1.**
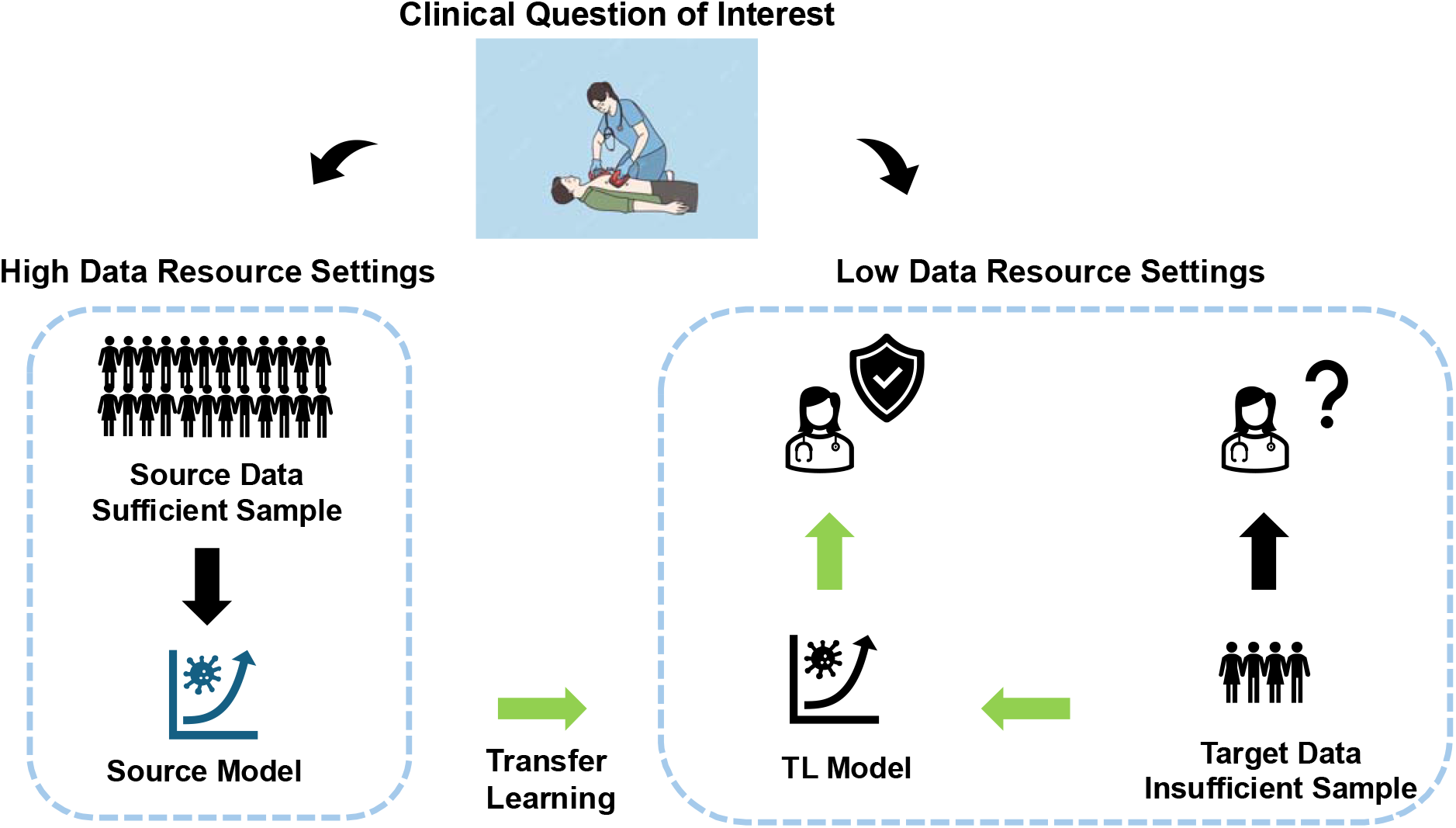
Illustration of the transfer learning (TL) approach. A source model, trained on large source data from high-resource settings, often performs poorly when applied to small target data from low-resource settings. A target model, trained only on target data, may also be suboptimal. TL improves predictive performance by adapting the source model using limited target data, producing a TL model better suited for low-data-resource settings (LDRS).

To demonstrate the applicability of TL, we aimed to address a clinical question requiring accurate risk assessment that is influenced by regional data variability due to epidemiological differences and other factors contributing to global healthcare disparities. Out-of-hospital cardiac arrest (OHCA) was chosen as a proof of concept, as it is a life-threatening emergency where the heart suddenly stops functioning outside a hospital, and immediate medical intervention is crucial. Oxygen deprivation to the brain and other vital organs can cause irreversible damage within minutes^13,14^. Even when return of spontaneous circulation (ROSC) is achieved and patients receive intensive care, many OHCA patients suffer from poor neurological outcomes due to ischemic injury sustained during the arrest^15,16^. Predicting neurological recovery is therefore useful for guiding clinical decisions, such as whether to continue intensive care or withdrawing life-sustaining treatment^15^. However, in LDRS, where data collection is particularly challenging, large-scale datasets for OHCA studies are often unavailable, complicating the development of accurate predictive models.

In response to these challenges, we applied TL to develop a neurological outcome prediction model specifically tailored for regions with limited data, using an existing external model and the Pan-Asian Resuscitation Outcomes Study (PAROS) network^17^. Our goal is to demonstrate TL’s capacity to improve predictive accuracy for critical outcomes, such as OHCA, and extend its applicability to other conditions in LDRS. Through this study, we aim to showcase how TL can drive global healthcare innovation by creating scalable, equitable AI models that can improve patient outcomes and help reduce healthcare disparities worldwide.

## Methods

### Study design and setting

This study was a secondary and retrospective analysis of data from the PAROS registry^17^. PAROS is a collaborative clinical research initiative established by international emergency medical professionals and researchers to investigate the epidemiology of OHCA across the Asia-Pacific region^17–19^. Specifically, this analysis utilized data from the PAROS 2 international dataset, a prospective, observational registry encompassing OHCA cases from 13 regions^20^.

To ensure uniformity in outcome reporting, all PAROS participants adhered to a standardized taxonomy and data collection protocol. The registry included all OHCA cases reported by emergency medical services (EMS), defined as the absence of a pulse, unresponsiveness, and apnea within the participating regions. A wide range of variables was collected, including patient demographics (e.g., age, gender), event-specific information (e.g., location type, bystander cardiopulmonary resuscitation (CPR)), and EMS-related data (e.g., drug administration).

### Study Population

This study analyzed adult OHCA patients recorded in the PAROS database from January 2017 to December 2021 in Singapore and Vietnam. Pediatric patients (<18 years), those missing epinephrine data, or those without detailed neurological outcome records were excluded. Two local cohorts were formed to represent settings with different sample sizes: a small sample size cohort from Vietnam (Ho Chi Minh City, Hue, Hanoi) and a large sample size cohort from Singapore.

### Variables

The variables selected for outcome predictions are consistent with those used in the Japan study^21^. These include: age (in years), gender (male/female), first recorded cardiac rhythm (ventricular fibrillation (VF)/pulseless ventricular tachycardia (pVT), pulseless electrical activity (PEA) or asystole), no flow time (duration between collapse and CPR start in minutes), low-flow time (time from initiation of CPR to return of spontaneous circulation (ROSC), in minutes), use of bystander automated external defibrillator (AED) (yes/no), prehospital defibrillation (yes/no), prehospital administration of epinephrine (yes/no) and cardiac rhythm at emergency department (ED) arrival (VF/pVT, PEA, asystole or ROSC). Details of these variables are available in Table S2 in the Supplementary Materials.

### Outcomes

The primary outcome was binary, defined as survival with favorable neurological outcomes (Cerebral Performance Category (CPC) score of 1 or 2) at 30 days post-arrest or at discharge. The CPC score was assessed by the treating physician.

### Data preprocessing

Missing values in the predictors were imputed using the missForest R package^22^, a nonparametric missing value imputation method widely used in clinical and biomedical studies^23,24^. For numerical predictors, additional data transformations were applied: age, no flow time, and low flow time were standardized, and a log transformation was applied to the low-flow time variable to align with the methodology of the Japanese study^21^.

### Prediction modeling

The original model developed by Nishioka et al.^21^ used least absolute shrinkage and selection operator (Lasso) regression to predict neurological outcomes in Japanese OHCA patients, achieving an AUROC of 0·943 (95% confidence interval (CI): 0·934–0·953). This model was trained on the Osaka CRITICAL database^21^ (17,385 patients) and validated using the JAAM-OHCA registry^21^ (29,633 patients), providing a robust foundation for neurological outcome prediction.

To adapt this model to the PAROS datasets, we employed the Trans-Lasso^25^ algorithm, which integrates TL techniques tailored for Lasso regression models. The Trans-Lasso algorithm was initialized with the model parameters reported in the original Japanese study and then refined using data from the PAROS registry to create cohort-specific predictive models. For baseline comparisons, the performance of the original Japanese model (external model) was evaluated directly on both the Vietnam and Singapore cohorts. The implementation of the Trans-Lasso algorithm used in this study can be accessed at https://github.com/nliulab/Clinical-Transfer-Learning.

The same predictors as the external model were used in our analysis, but the outcome of interest was redefined as good neurological outcomes. Model performance was assessed using the AUROC and the area under the precision-recall curve (AUPRC). Sensitivity and specificity values were also calculated to evaluate the model’s ability to correctly identify cases with and without good neurological outcomes. All statistical analyses were conducted using R software version 4.3.1 (The R Foundation for Statistical Computing, Vienna, Austria).

### Ethical statement

This retrospective analysis was approved by the relevant ethics committees at each participating PAROS site and by the Centralized Institutional Review Board and Domain Specific Review Board for Singapore (reference numbers: 2013/604/C, 2013/00929 and 2018/2937). Informed consent was waived due to the observational nature of the study, and all data were de-identified.

## Results

The Vietnam and Singapore cohorts consisted of 243 and 15,916 patients, respectively. Figure 2 illustrates the cohort formation process, with a 6:4 split between training and testing datasets for both cohorts. Details on missing data proportions are available in Supplementary Table S1. The descriptive statistics are summarized in Table 1, with continuous variables presented as medians with interquartile ranges, and categorical variables reported as counts and percentages. Compared to the original Japanese study cohort, the Vietnam dataset featured significantly younger patients, contributing to greater data heterogeneity.

**Table 1.** Description of the study cohorts. Data are presented as count (percentage) of patients unless otherwise indicated.

|  |  | Vietnam (n = 243) | Singapore (n = 15,916) |
| --- | --- | --- | --- |
| Age (years), median (Q1-Q3) |  | 54 (42-67) | 72 (59-83) |
| Gender (Male) |  | 186 (76.5) | 10,058 (63.2) |
| Bystander AED (Yes) |  | 10 (4.0) | 1364 (8.6) |
| First Rhythm, n(%) | VF/pVT | 38 (15.6) | 2273 (14.3) |
|  | PEA | 0 (0) | 4391 (27.6) |
|  | Asystole | 205 (84.4) | 9252 (58.1) |
| No Flow Time (min), median (Q1-Q3) |  | — | 14 (5-22) |
| Low Flow Time (min), median (Q1-Q3) |  | — | 42 (39-50) |
| Prehospital Defibrillation (Yes) |  | 38 (15.6) | 3281 (20.6) |
| Prehospital Epinephrine (Yes) |  | 111 (45.7) | 12,197 (76.6) |
| Prehospital Advanced Airway (Yes) |  | 100 (41.2) | 13,966 (87.7) |
| Rhythm at ED arrival, n(%) | VF/pVT | 19 (7.8) | 1580 (9.9) |
|  | PEA | 1 (0.4) | 1959 (12.3) |
|  | Asystole | 147 (60.5) | 10,622 (66.7) |
|  | ROSC | 76 (31.3) | 1755 (11.0) |
| Neurological Outcome (Good) |  | 13 (5.2) | 572 (3.6) |

AED, Automated external defibrillator, VF, ventricular fibrillation, VT, ventricular tachycardia, PEA, pulseless electrical activity, ROSC, return of spontaneous circulation, ED, emergency department. Q1, 25%tile, Q3, 75%tile.
Variables 'No Flow Time' and 'Low flow time' are not available in the Vietnam dataset, and their descriptions are reported only for the Singapore dataset.

**Figure 2.**
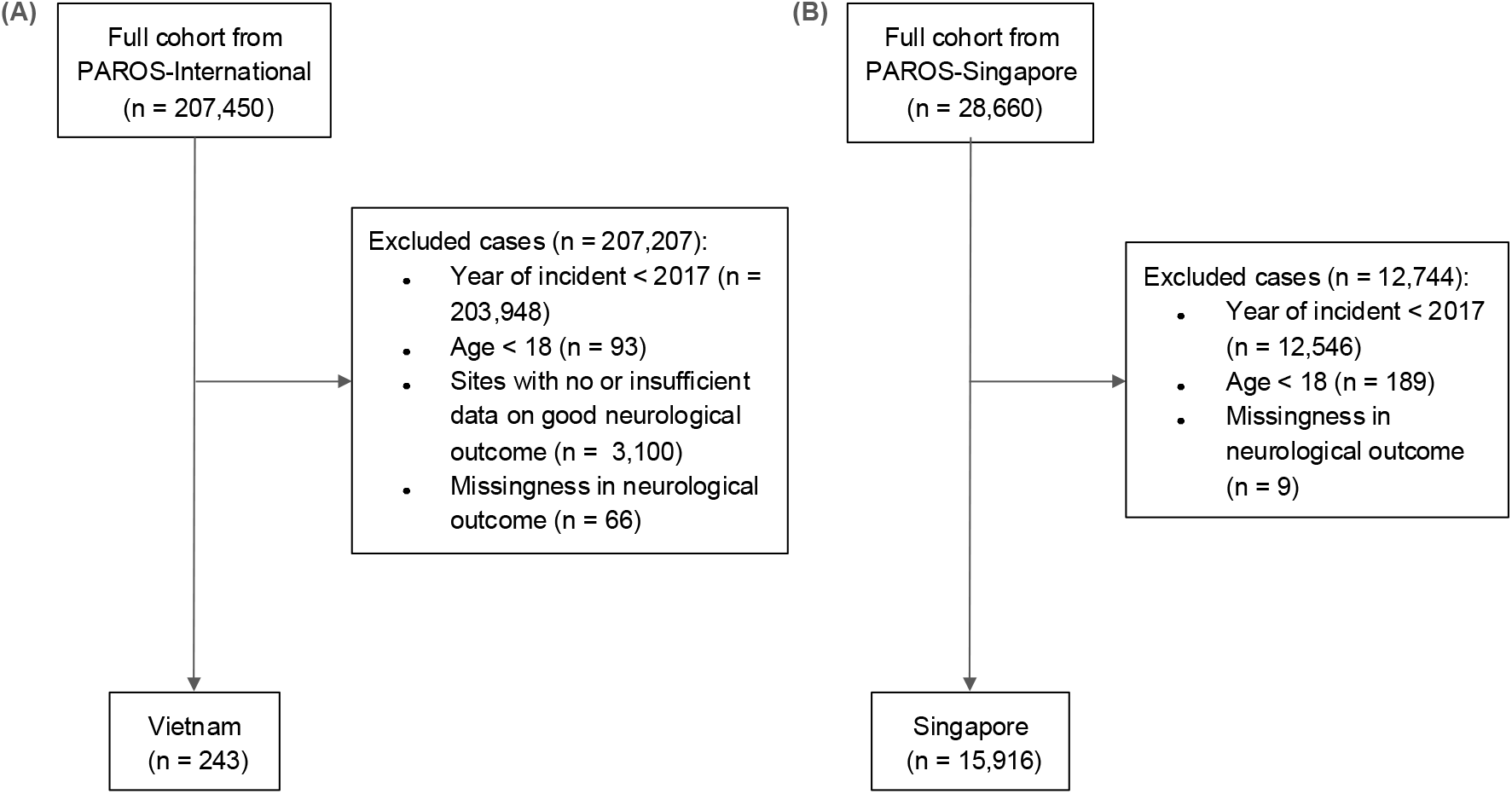
Flowchart for cohort formation.

TL models were independently fitted using the training datasets for Vietnam and Singapore and evaluated on their respective testing datasets and compared with the external model. As shown in Table 2, the TL model significantly outperformed the external model in both cohorts. The TL-Vietnam model, adapted using local data from Vietnam, improved performance compared to the external model, achieving an AUROC of 0·807 (95% CI: 0·626–0·948), up from the external model’s AUROC of 0·467 (95% CI: 0·141–0·785). Similarly, the TL-Singapore model, adapted using the local data from Singapore, achieved an AUROC of 0·955 (95% CI: 0·940–0·967), slightly outperforming the external model’s AUROC of 0·945 (95% CI: 0·929–0·958).

**Table 2.** Comparison of model performance.

| **Cohort** | **Model Type** | **AUROC (95% CI)** | **AUPRC** | **Specificity at Sensitivity = 0·55** | **Specificity at Sensitivity = 0·85** |
| --- | --- | --- | --- | --- | --- |
| Vietnam (n=243) | Source | 0·467 (0·141-0·785) | 0·428 | 0·707 | 0·000 |
|  | TL | 0·807 (0·626-0·959) | 0·889 | 0·902 | 0·511 |
| Singapore (n=15916) | Source | 0·945 (0·929-0·958) | 0·527 | 0·986 | 0·908 |
|  | TL | 0·955 (0·940-0·967) | 0·885 | 0·990 | 0·920 |

AUROC, Area under the receiver operating characteristic curve, AUPRC, Area under the precision-recall curve, CI, Confidence interval, TL, Transfer learning

The AUPRC also improved with the application of TL. On the Vietnam dataset, AUPRC increased from 0·428 for the external model to 0·889 for the TL-Vietnam model. On the Singapore dataset, AUPRC increased from 0·527 to 0·885. Specificity values at fixed sensitivity thresholds, detailed in Table 2, further confirmed that the TL models consistently outperformed the external model.

The parameters of the TL-Vietnam and TL-Singapore models are provided in Supplementary Table S2. These findings demonstrate the ability of TL to enhance model performance, particularly in adapting externally developed models to diverse geographic and resource-limited settings.

## Discussion

This study demonstrates the feasibility and utility of TL in adapting existing predictive models to new clinical contexts, especially in LDRS. Using OHCA as a proof of concept, we showed that TL can significantly enhance model performance with smaller datasets, as observed in Vietnam, and provide incremental benefits with larger datasets, as seen in Singapore. These findings highlight TL’s broad applicability across diverse global healthcare settings, offering a scalable solution for developing robust predictive models and reducing healthcare disparities by tailoring models to specific regional contexts.

Healthcare research in LDRS faces numerous challenges, including small population sizes, low outcome prevalence, and disparities in epidemiological features and data collection systems. Importantly, even high-income regions are not immune to these issues. For example, in OHCA, where favorable neurological outcomes are rare (3–4%), large datasets are essential for robust model development. The external model used in this study was developed from 46,918 OHCA patients in Japan over 5–6 years^21^, providing a solid foundation for model development and validation. In contrast, Singapore, with only approximately 3,000 annual OHCA cases, would require more than a decade to accumulate a comparable dataset. While both Singapore and Japan are considered developed countries, conducting such research in Singapore is more challenging due to differences in population size. The situation is even more difficult in data-constrained settings like Vietnam, where OHCA databases are still emerging. Despite approximately 4,000 annual cases, it would still take over 10 years to collect enough data for robust model development. These prolonged timelines delay the implementation of predictive models that could improve patient outcomes and exacerbate global healthcare disparities, highlighting the urgent need for innovative solutions to address data gaps in LDRS.

AI techniques like TL provide an effective solution by adapting existing models to local populations, improving predictive performance even with scarce data. This study highlights the transformative potential of TL for developing accurate prediction models in LDRS, where socioecological factors, in addition to financial constraints^3^, influence healthcare outcomes. In Vietnam (n = 243), TL significantly enhanced model accuracy. In contrast, in Singapore (n = 15,916), TL provided more modest gains, illustrating its adaptability across different resource levels. Despite Singapore’s larger dataset, the annual OHCA case numbers remain insufficient to independently develop models comparable to those built with Japan’s extensive data. These findings underscore the utility of TL in leveraging external information to overcome local dataset limitations, enabling the development of clinically relevant models tailored to specific populations. By addressing heterogeneity in data distributions and clinical practices, TL enhances the reliability and applicability of predictive models, making AI technologies more accessible and impactful in data-limited settings.

As shown in Table 2, the benefits of TL were most pronounced with the Vietnam cohort, where the improvement in overall prediction performance was more significant than in Singapore. This supports the theoretical foundations of TL^10^, which is particularly effective when the source data (e.g., the Japanese cohort) has a large sample size and the target data (e.g., Vietnam and Singapore) is relatively limited. This study demonstrates the applicability of TL in challenging settings like LDRS, showing how researchers can directly leverage knowledge from external studies without requiring access to additional datasets or direct collaborations. Federated learning (FL), another relevant AI technique, addresses data privacy concerns in cross-site collaboration by enabling co-training of AI models without sharing data^26^. Unlike FL, which requires simultaneous participation from multiple data owners, TL allows independent adaptation of publicly available models. This makes TL particularly advantageous for researchers with limited access to external data or collaboration opportunities, offering a practical way to overcome barriers associated with data sharing and resource constraints^10^.

TL is a flexible AI approach applicable to a wide range of models and knowledge transfer scenarios^10^, such as transferring insights on drug sensitivity between different cancer types^27^ or adapting knowledge of surgical complications across patient groups^28^. In LDRS, where collecting large, high-quality datasets is challenging, TL offers an efficient way to leverage existing models for improved predictive performance. Regardless of the clinical or biomedical domain, as Li et al.^10^ highlight, the successful implementation of TL depends on carefully selecting an appropriate external study and ensuring that the chosen TL framework aligns with the research question. It is also crucial to address potential privacy concerns when building or using external models. When co-training is required, FL can be integrated into TL frameworks to enable secure collaborations across multiple sites, ensuring data privacy while still facilitating knowledge transfer.

One reason for TL’s effectiveness in this study may be the standardized data format and consistent variable definitions under the PAROS study framework. The Utstein data format, adopted in this study, has been globally accepted since its establishment in 1995, and most countries utilize it for OHCA data collection^29^. Furthermore, the prehospital management pathways for OHCA cases follow similar protocols across the study countries—paramedics respond to an emergency call, perform resuscitation, and transport patients to hospitals. During this clinical pathway, prehospital OHCA data are systematically collected, which likely facilitated effective knowledge transfer using TL.

Another potential reason is that certain predictors, such as ROSC status, are expected to be strongly associated with patient outcomes, despite variability in unmeasured clinical factors like in-hospital care or hospital systems among the three countries^30^. Given these considerations, further research is essential to investigate the applicability of TL to the other clinical scenarios where unmeasured factors may significantly impact outcomes. For example, in patients with refractory VF, treatment strategies vary markedly across countries. In Japan, advanced invasive resuscitation procedures, such as extracorporeal cardiopulmonary resuscitation, are commonly performed and may contribute improved outcomes^30^. In contrast, such procedures are rarely utilized in Singapore and Vietnam^30^. It remains unclear whether TL would perform well in contexts where critical unmeasured factors, such as the treatment strategy or in-hospital care, vary substantially and may significantly affect patients’ outcomes.

Future research should explore strategies for handling ultra-small datasets, where outcome prevalence is extremely low or absent. In this study, the Vietnam dataset included 234 cases with only 13 positive outcomes, but settings with even fewer cases present greater challenges. In such instances, the TL method used here may not be directly applicable. To enhance model robustness and improve predictive reliability, alternative approaches—such as synthetic data generation and oversampling—should be integrated with TL. Combining these techniques within TL frameworks may further mitigate data scarcity in low-resource clinical settings.

This study has several limitations. First, while the Singapore data were derived from a nationwide population-based registry for OHCA patients and are likely representative of the Singapore population, the Vietnam data were collected from a limited number of hospitals in Ho Chi Minh, Hanoi and Hue. As a result, the Vietnam data may not be representative of the broader population, potentially limiting the generalizability of the model to current clinical settings in Vietnam. Second, although the data from Vietnam, Singapore, and Japan were collected using an internationally standardized format with consistent definition, differences in local clinical practices and resuscitation protocols may have influenced data collection and reporting. This might lead to a risk of measurement bias. Third, prospective external validation using independent data would be ideal for confirming the robustness of our results. However, at present, no such dataset is available in Vietnam or Singapore for this process. Further research is warranted to assess reproducibility and validate these findings in independent cohorts.

In conclusion, this study demonstrates that TL has the potential to bridge regional resource disparities in healthcare. While applied here to OHCA, TL’s adaptability extends to other emergency conditions with low event rates, such as trauma, sepsis, heart failure, and stroke. Moreover, TL holds broader potential beyond emergency medicine, offering a scalable, efficient strategy to enhance clinical decision-making in LDRS globally. By reducing reliance on large-scale data collection, TL facilitates equitable global access to AI-driven predictive models, helping to reduce healthcare disparities and optimize patient outcomes in data-constrained environments.

## Data Availability

The data supporting the findings of this study are not publicly available due to privacy and institutional data-sharing agreements.

## Acknowledgement

We would like to express our gratitude to all the other participating members of the Pan-Asian Resuscitation Outcomes Study Clinical Research Network (PAROS CRN) for their invaluable contributions. Participating Site Investigators: H Tanaka (Kokushikan University); SD Shin (Seoul National University College of Medicine); MHM Ma (National Taiwan University Hospital Yunlin Branch); K Kajino (Kansai Medical University); CH Lin (National Cheng Kung University); CW Kuo (Chang-Gung Memorial Hospital); S Karim (Hospital Sungai Buloh); S Jirapong (Rajavithi Hospital); P Khruekarnchana (Rajavithi Hospital); RH Ho (Chonnam National University Medical School and Hospital); HW Ryoo (Kyungpook National University); Tagashi Tagami (Nippon Medical School Tama Nagayama Hospital); PCI Ko (National Taiwan University, Yun-Lin Branch Hospital); KD Wong (Hospital Pulau Pinang); N Sumetchotimaytha (Rajavithi Hospital); FJ Gaerlan (Southern Philippines Medical Center); B Velasco (East Avenue Medical Center), GV Ramana Rao (GVK Emergency Management and Research Institute Telangana); W Cai (Zhejiang Provincial People’s Hospital); S Fei (Beijing Chaoyang Hospital); N Khan (Aga Khan University); ME Sayed (American University of Beirut Medical Center); MI Abuamllouh (Dubai Corporation for Ambulance Services); CB Yang (Sarawak General Hospital); Vimal M (GVK Emergency Management and Research Institute); Rajanarsing Rao HV (GVK Emergency Management and Research Institute); M Khursheed (National Institute of Cardiovascular Diseases); PJ Tiglao (Corazon Locsin Montelibano Memorial Regional Hospital); Zhou SA (Zhejiang Provincial People’s Hospital); A Alhumodi (Abu Dhabi Police GHQ).

We would like to thank Ms Pin Pin Pek and Ms Nur Shahidah from the Prehospital and Emergency Care Research Centre, Duke-NUS Medical School for coordination of the study, and Ms Patricia Tay from the Singapore Clinical Research Institute for her role as Network Secretariat for the PAROS CRN.

We also extend our sincere appreciation to Dr. Chuan Hong (Duke University) and Dr. Molei Liu (Peking University) for their invaluable expertise, support, and guidance in providing the source code for implementing the Trans-Lasso algorithm, which significantly contributed to the analytic framework of this study.

## Authors contribution

**Siqi Li**: Conceptualization, Methodology, Analysis, Project administration, Writing – original draft, Writing – review & editing. **Yohei Okada**: Methodology, Analysis, Data curation, Writing – original draft, Writing – review & editing. **Wenjun Gu**: Analysis, Data curation, Writing – original draft, Writing – review & editing. **Michael Hao Chen**: Data curation, Writing – review & editing. **Son Ngoc Do**: Data acquisition, Writing – review & editing. **Pham Dinh Quyet**: Data acquisition, Writing – review & editing. **Quoc TA Hoang**: Data acquisition, Writing – review & editing. **Marcus Eng Hock Ong**: Investigation, Resources, Writing – review & editing. **Nan Liu**: Conceptualization, Project administration, Funding acquisition, Resources, Writing – review & editing. All authors agree to be accountable for all aspects of the work.

## Supplementary Materials

**Table S1.** Missing data rates for predictors in the Vietnam (n=243) and Singapore (n=15,916) cohorts, presented as counts and percentages. Variables ‘No Flow Time’ and ‘Low flow time’ are not available in the Vietnam dataset, and their missing rates are reported only for the Singapore dataset.

| Predictors | Missing Rate, n (%) |  |
| --- | --- | --- |
|  | Vietnam (n=243) | Singapore (n=15,916) |
| Age | 0 (0) | 0 (0) |
| Gender | 0 (0) | 0 (0) |
| Bystander AED | 79 (32.5) | 0 (0) |
| First rhythm | 224 (92.2) | 1179 (7.4) |
| No flow time | — | 9759 (61.3) |
| Low flow time | — | 12290 (77.2) |
| Prehospital defibrillation | 167 (68.7) | 0 (0) |
| Prehospital epinephrine | 168 (69.1) | 7580 (47.6) |
| Prehospital advanced airway | 167 (68.7) | 58 (0.4) |
| Rhythm at ED arrival | 4 (1.6) | 15103 (94.9) |

AED, Automated external defibrillator, ED, emergency department.

**Table S2.**
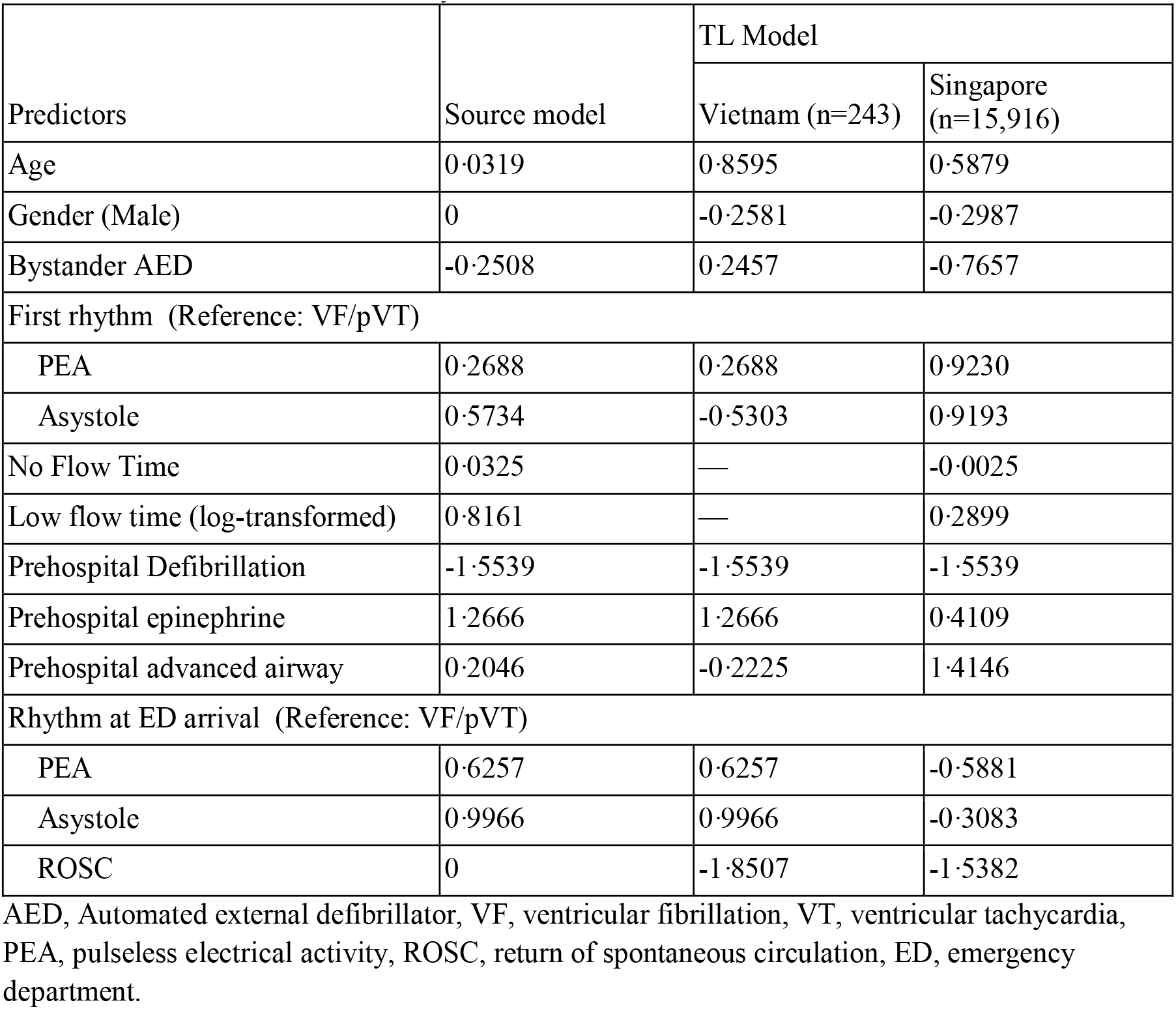
Lasso regression model coefficients for the source and transfer learning (TL) models applied to cardiac arrest datasets. Variables ‘No Flow Time’ and ‘Low Flow Time’ are unavailable in the Vietnam dataset, as indicated by the omission of coefficients.

**Table S3.** Further details for definition of the data.

| **Variable** | **Definition/Explanation** |
| --- | --- |
| Age | Patient age |
| Gender | Patient gender (Male/Female) |
| Bystander AED | Shock provided with automated external defibrillator (AED) by lay person |
| First rhythm | Initial cardiac rhythm confirmed by paramedics (ventricular fibrillation[VF] or ventricular tachycardia [VT]/Pulseless electrical activity [PEA]/Asystole) |
| No flow time | Duration between collapse and CPR start (minute) |
| Low flow time | Duration between CPR start and return of spontaneous circulation [ROSC] |
| Prehospital defibrillation | Shock provided by paramedics during resuscitation |
| Prehospital epinephrine | Administration of epinephrine by paramedics during resuscitation |
| Prehospital advanced airway | Insertion of advanced airway (intubation/supraglottic airway device) by paramedics |
| Rhythm at ED arrival | Initial cardiac rhythm at emergency department (ED) arrival (ventricular fibrillation[VF] or ventricular tachycardia [VT]/Pulseless electrical activity [PEA]/Asystole/ROSC) |
| Neurological outcome | Cerebral performance category 1 or 2 at hospital discharge or 30-day after the cardiac arrest |

